# Characterisation of *Staphylococcus argenteus* carried by healthy Royal Marines: a molecular epidemiology case-study

**DOI:** 10.1101/2021.06.03.21257959

**Authors:** Elita Jauneikaite, Bruno Pichon, Mia Mosavie, Joanne L. Fallowfield, Trish Davey, Neil Thorpe, Andrew Nelstrop, Shiranee Sriskandan, Lucy E Lamb

**Author notes:** **Corresponding authors**: Dr Elita Jauneikaite, Imperial College Research Fellow, Imperial College London, London W2 1PG, UK. E-mail address, Dr Lucy E Lamb, Defence Consultant Infectious Diseases and General Medicine, Royal Free NHS Foundation Trust, London NW3 2QG, UK.

## Abstract

**Objectives:** During a prospective study of *S. aureus* carriage in Royal Marines (RM) recruits, six *S. argenteus* strains were identified in four recruits undertaking military training together. As *S. argenteus* sepsis leads to mortality similar to *S. aureus*, we determined the potential for person-to-person transmission, to evaluate future outbreak risk.

**Methods:** We used whole-genome sequencing to characterise *S. argenteus* and investigate phylogenetic relationships between isolates. Participant colonisation with *S. aureus* and skin and soft tissue infection acquisition were recorded.

**Results:** All six *S. argenteus* strains were *spa*-type t5078, ST2250. Strains were detected in 4/40 recruits in the same troop (training cohort) in weeks 1, 6 or 15 of training. No *mec, tsst* or *LukPV* genes were detected. We identified differences of 10-35 core SNPs between *S. argenteus* from different recruits. In two recruits, two *S. argenteus* strains were isolated; these could be distinguished by 3 and 15 core SNPs in each case. *S. argenteus* was not identified in any one of the other 21 participating troops (1,012 recruits).

**Conclusions:** The identification of *S. argenteus* within a single troop from the total recruit population supports a common source for transmission, supported by SNP analysis. The high number of SNPs between some isolates may indicate a common source of diverse isolates or a high level of *S. argenteus* mutation in carriage. *S. argenteus* ST2250 is a newly recognised lineage; a better understanding of the frequency of genetic changes during transmission and transition from asymptomatic carriage to disease is required.

## Introduction

*Staphylococcus argenteus* is a newly identified staphylococcus species, previously known as *S. aureus* clonal complex 75 [1]. Globally, it is known to be a sporadic pathogen causing sepsis [2], skin and soft tissue infections [3] and bone infections [3,4]. Data on carriage and potential person-to-person transmission are scarce: two studies report carriage rates of 0.8% (5/563) and 3.4% (5/146) of *S. argenteus* in healthy individuals [5,6], while just one study has described familial transmission of *S. argenteus* [7]. Detailed genomic analysis currently has been confined to the disease-causing *S. argenteus* populations in Asia [8] and Denmark [9]. There is presently no information on the spread and genomic features of carried *S. argenteus* strains in the UK.

Within a large prospective observational study of *S. aureus* carriage in Royal Marines (RM) recruits in the UK (n=1012), six *S. argenteus* isolates were identified [10]. To clarify the role of person-to-person transmission, we investigated the clustering of these isolates in relation to the time, location, and disease acquisition data of participants and undertook whole genome sequencing of these strains. We evaluated the genotypes, antibiotic resistance and virulence genes maintained by the six *S. argenteus* strains and compared these with available *S. argenteus* genomes collected globally.

## Materials and Methods

### Asymptomatic carriage cohort study

The longitudinal nasal swab sampling of staphylococcal isolates from RM recruits in training has been previously described [10]. The study was approved by the Ministry of Defence Research Ethics Committee (MODREC 172/Gen/10).

### Bacterial samples, genomic sequencing and analysis

Genomic DNA of confirmed *S. argenteus* isolates was extracted using an adapted protocol (Supplementary Methods). DNA libraries were prepared with the Nextera XT kit (Illumina, Cambridge, UK) and sequenced on the Illumina HiSeq 2500 instrument (Illumina), generating 100 base paired end fragments. Raw reads were *de novo* assembled and annotated; multi-locus sequence type (MLST), antibiotic resistance genes, virulence genes, plasmids and phage information were derived as described in Supplementary Methods. Phylogenetic relationships between *S. argenteus* strains were characterised by building a phylogenetic tree from extracted core SNPs mapped against study isolate Sarg_A1 (Supplementary Methods). The sequence data have been submitted to the European Nucleotide Archive, under the study accession PRJEB41767 (Table S1).

## Results

### Epidemiology

*S. argenteus* was detected in recruit D at the start of training, but not in any other of the 1011 recruits, yielding a background asymptomatic carriage rate of 0.09%. Within recruit D’s troop, by week 6 of training, another two recruits demonstrated *S. argenteus* and a fourth recruit demonstrated *S. argenteus* carriage by week 15 (Table S2). Over the entire sampling period *S. argenteus* was found only in these four out of 1012 recruits (study prevalence 0.4%), and not in any other troop undertaking military training at the same time.

### Genomic characterisation of *S. argenteus* strains

The six isolates were multi-locus sequence type ST2250 (Table 1) and susceptible to erythromycin, clindamycin, mupirocin, cefoxitin and oxacillin by disk diffusion. They did not carry the Staphylococcal Chromosomal Cassette (SCCmec) harbouring the *mec* genes, but had *blaZ, aph(3’)-III, fosB* and *tet(L)* antibiotic resistance genes present (Table 1); no known AMR associated chromosomal mutations were detected (Table S3). No superantigenic enterotoxins associated with toxic shock (i.e., *tsst, sea, seb, sec, eta*, or *LukPV* genes) were identified in any of the *S. argenteus* strains. However, other major staphylococcal virulence genes were present (Table 1). Further genome interrogation showed presence of a 23,892–23,914bp^*^ long plasmid, that had *rep5, rep16*, and *blaZ* genes present on the *Tn552* transposon, and metal resistance factors *arc3, cadC* and *czcD* (Figure S1). Two prophages (one intact (∼45.5-58.7kb), one incomplete (∼5.9kb)) were present encoding leukocidins *LukDv* and *LukEv*, two enterotoxin-like genes, and the *sak* gene.

**Table 1.** Summary of characteristics of *S. argenteus* strains.

| Isolate ID | RM recruit | Week of isolation | Swab | MLST | Acquired antimicrobial resistance genes |  |  |  | Virulence genes |  |  |  |  |  |
| --- | --- | --- | --- | --- | --- | --- | --- | --- | --- | --- | --- | --- | --- | --- |
|  |  |  |  |  | B-lactams | Aminoglycosides | Tetracycline | Fosfomycin | *coa* | *sak* | *scn* | *hly/hla* | *hlgABC* | *lukED* |
| Sarg_A1 | A | 6 | Throat | ST2250 | *blaZ* | *aph(3')-III* | *tet(L)* | *fosB* | y | y | y | y | y | y |
| Sarg_B1 | B | 15 | Throat | ST2250 | *blaZ* | *aph(3')-III* | *tet(L)* | *fosB* | y | y | y | y | y | y |
| Sarg_C1 | C | 6 | Nose | ST2250 | *blaZ* | *aph(3')-III* | *tet(L)* | *fosB* | y | y | y | y | y | y |
| Sarg_C2 | C | 6 | Throat | ST2250 | *blaZ* | *aph(3')-III* | *tet(L)* | *fosB* | y | y | y | y | y | y |
| Sarg_D1 | D | 1 | Nose | ST2250 | *blaZ* | *aph(3')-III* | *tet(L)* | *fosB* | y | y | y | y | y | y |
| Sarg_D2 | D | 1 | Throat | ST2250 | *blaZ* | *aph(3')-III* | *tet(L)* | *fosB* | y | y | y | y | y | y |

### Person-to-person transmission and comparison to global *S. argenteus* genomes

The six *S. argenteus* genomes were compared to a reference genome Sarg_A1 to assess strain diversity. Pair-wise core SNPs analysis showed that genetic diversity of these six strains ranged from 3–35 SNPs (Table S3). Sequences Sarg_C1 and Sarg_C2 differed by 3 SNPs; a level of low diversity that would be expected, as both strains were obtained from recruit C (Table 1). However, Sarg_D1 and Sarg_D2, which were both from recruit D (Table 1), differed by 15 SNPs (Table S4).

Phylogenetic comparison with global ST2250 *S. argenteus* strains indicated that the six isolates were 52–56 SNPs different from the nearest *S. argenteus* isolate from Thailand (Figure 1). The distinct clustering of our six isolates in the global tree excludes the possibility of multiple introductions of the lineage and suggests dispersion of this variant within the troop. Pangenome analysis showed 2,271 core genes were shared by 124 ST2250 *S. argenteus* isolates and no genes were found to be unique only to the six isolates in our study.

**Figure 1.**
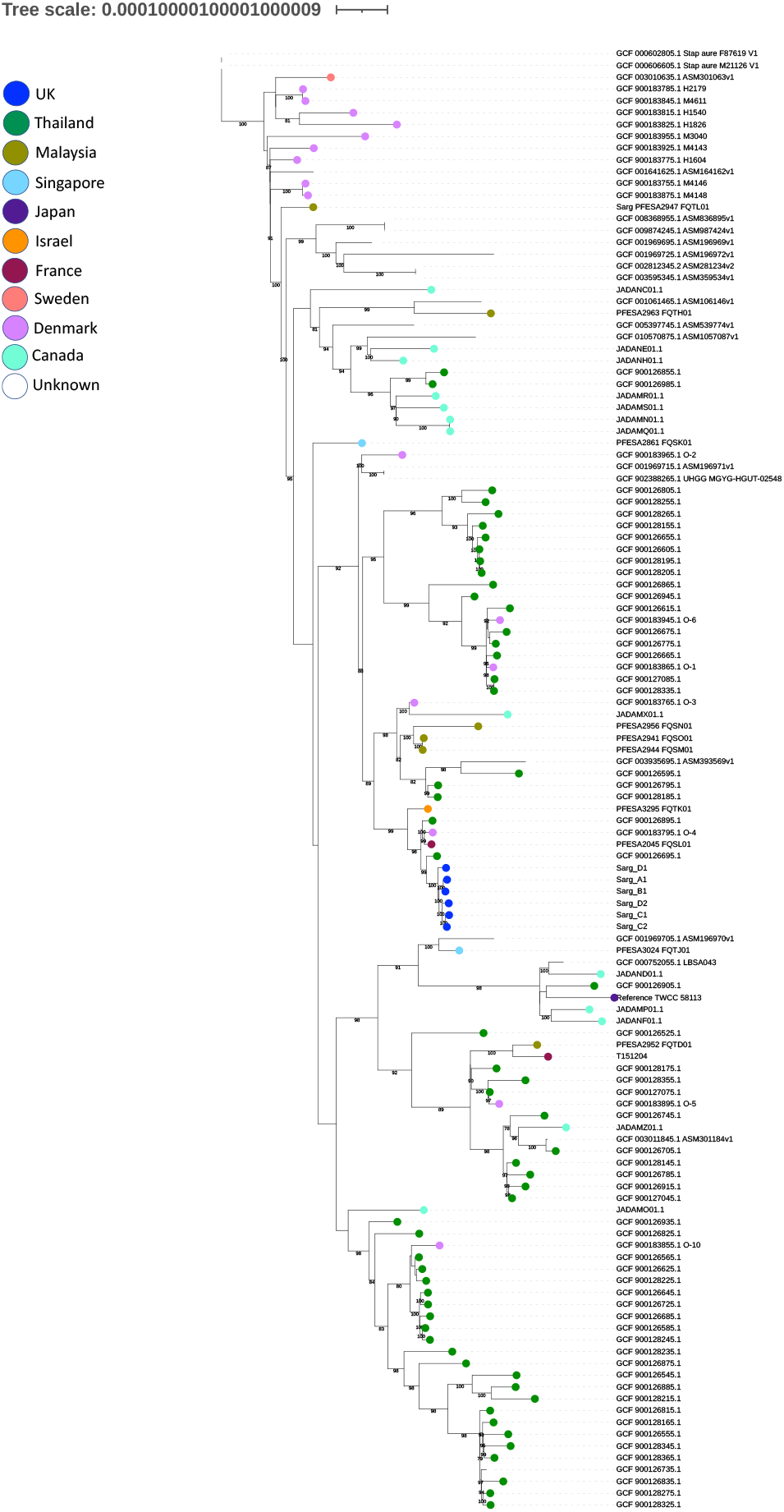
Phylogenetic relationship between global *S. argenteus* ST2250 including the six *S. argenteus* strains investigated in this study. Maximum-likelihood phylogenetic tree constructed from full alignments (including gaps, Ns and invariant sites) of global ST2250 *S. argenteus* genomes (n=118, Table S5) and 6 genomes from this study using reference *S. argenteus* TWCC 58113 (acc. no. AP018562.1) using IQtree with bootstrapping of 1000 replicates (see Suppl. Methods for more details). The six strains from RM recruits (coloured in blue) cluster together, no other *S. argenteus* ST2250 genomes were available from the UK at the time. Nearest strain was ST2250 from Thailand with 52-56 SNPs different from the *S. argenteus* strains analysed in this study, followed by a cluster of 3 strains (from France, Denmark and Thailand) with 64-72 SNPs. Colour of the circle at the end of the branch indicate the country of *S. argenteus* origin, no circle means the information on country of origin was not available. The scale bar indicates nucleotide substitutions per site.

## Discussion

This study describes a cluster of *S. argenteus* carriage strains in a small group of RM recruits during training, pointing strongly to potential transmission events within the troop or presence of a common source. Genomic analysis demonstrated 3–35 SNPs difference between individual isolates, from which it might be construed that the isolates were not related. However, it is proposed that this is highly unlikely because: (i) the cluster was limited to a small number of recruits who lived and trained together and was not identified in any other of 1,012 recruits from the longitudinal study [10]; (ii) even within-host, changes of 15–19 whole-genome SNPs have been observed in MRSA, pointing to a high within-host mutation rate, resulting in cloud diversity following long term carriage [11,12]. Thresholds of 25 whole-genome SNPs or 15 core-genome SNPs have been suggested for inferring person-to-person transmission of MRSA within 6 months [12].

Our *S. argenteus* isolates were methicillin-susceptible but showed a multidrug resistance gene profile. Notably all six strains had *BlaZ, aph(3’)-III* and *tet(L*) genes, conferring resistance to beta-lactams, aminoglycosides and tetracyclines; these genes were also previously reported in other ST2250 *S. argenteus* strains [8,9]. *S. argenteus* has been reported to cause skin and soft infections but there is no consensus yet on whether *S argenteus*-associated cases are more severe than *S. aureus*-associated skin infections. Given the similar clinical manifestations caused by *S. aureus* and *S. argenteus*, the European Society of Clinical Microbiology and Infectious Diseases (ESCMID) Study Group for Staphylococci and Staphylococcal Diseases has suggested that the two should be distinguished when possible for research purposes, but there is no need to differentiate for diagnostic purposes [13].

In conclusion, *S. argenteus* (ST2250) was carried asymptomatically and transmitted between four healthy RM recruits in a large prospective population study of staphylococcal carriage. Although the *S. argenteus* isolates identified herein were not disease-associated and were methicillin-susceptible, the co-carriage of extracellular toxins and propensity for transmission underlines the need for further understanding of carriage patterns, within host diversity, and toxin production of ST2250 and other clones of *S. argenteus*. Future detailed investigation and surveillance of the clinical manifestations and outcomes of *S. argenteus* infections are required.

## Data Availability

The sequence data have been submitted to the European Nucleotide Archive, under the study accession PRJEB41767.

## Acknowledgement

The authors thank the CTCRM Medical Centre for support with sample collection. This study was funded by the Surgeon General’s Research Strategy Group, Ministry of Defence. EJ, MM, BP, SS and LEL acknowledge support from the National Institute for Health Research Health Protection Research Unit (NIHR HPRU) in Healthcare Associated Infections and Antimicrobial Infections in partnership with Public Health England (PHE), in collaboration with, Imperial Healthcare Partners, University of Cambridge and University of Warwick. SS acknowledges the NIHR Imperial Biomedical Research Centre. The views expressed in this publication are those of the author(s) and not necessarily those of the NHS, the National Institute for Health Research, the Department of Health and Social Care or Public Health England. EJ is an Imperial College Research Fellow, funded by Rosetrees Trust and the Stoneygate Trust.

## Author contributions

EJ, LEL conceptualization of the study. LEL acquired the funding and coordinated the project. EJ, LEL, PB curated the data. EJ and PB carried out bioinformatics analysis. MM, PB and EJ carried out laboratory work. SS, LEL supervised the team. SS, PB, JLF, TD, NT, AN and LEL provided study materials and accompanying demographic data for the samples. EJ prepared tables and figures. EJ, LEL wrote the original draft. All co-authors reviewed and edited further drafts.

## Footnotes

* size variation could be due to potential assembly mismatches of small repetitive regions within the putative plasmid.

