## Supplementary materials for "Characterisation of *Staphylococcus argenteus* carried by healthy Royal Marines: a molecular epidemiology case-study"

### Characterisation of *S. argenteus* carried between healthy Royal Marines: a molecular epidemiology case-study to track transmission

Elita Jauneikaite[1,2*], Bruno Pichon[2,3], Mia Mosavie[2], Joanne L. Fallowfield[4], Trish Davey[4], Neil Thorpe[4], Andrew Nelstrop[5,6], Shiranee Sriskandan[2] and Lucy E Lamb[2,7,8*]

### Supplementary Methods

**Antimicrobial susceptibility testing.** Antimicrobial susceptibility testing to erythromycin, clindamycin, mupirocin, cefoxitin and oxacillin was carried out by disk diffusion. The results were interpreted in accordance with the British Society for Antimicrobial Chemotherapy guidelines (http://bsac.org.uk/wp-content/uploads/2012/02/Version-12-Apr-2013_final.pdf).

***S. argenteus* DNA extraction.** The isolates were regrown on Columbia agar with 5% sheep blood plates (BD GmbH, Heidelberg, Germany) and incubated at 37°C overnight. Isolates were confirmed to be *S. argenteus* using the MALDI-TOF (MALDI Biotyper®, Bruker Daltonik GmbH, Germany). Overnight growth was resuspended in 100 µl of lysis buffer (100 mM NaCl, 10 mM Tris–HCl pH8, 1 mM EDTA, 1% Triton-X-100) with 20 µl of lysostaphin (1 mg/ml, Sigma, UK) and 20 µl of lysozyme (100mg/ml, Sigma, UK) incubated for 37 °C for 15 min before boiling for 10 min. Samples were centrifuged 13,000 × *g* for 2 min and the DNA-containing supernatant was further purified using an equal volume of chloroform. DNA was precipitated with isopropanol and resuspended in dd$H_2O$.

***Genomic analyses***

**Raw read assembly and annotation.**

Raw sequencing reads were checked using FastQC v.0.11.2 (https://www.bioinformatics.babraham.ac.uk/projects/fastqc/). Reads were then *de novo* assembled using SPAdes v. 3.10.1 [1], assembly statistics was checked using Quast v.4.5 [2]. Assemblies were annotated with Prokka v.1.13 [3].

**Genomic characterisation of *S. argenteus* genomes.**

SRST2 v0.2.0 [4] was used to determine Multi-Locus sequence type (MLST) using *S. aureus* MLST database, and staphylococcal virulence factors (as per Virulence Factors of Pathogenic Bacteria database, http://www.mgc.ac.cn/VFs/), identified potential virulence factors were then double checked with blasting sequences against NCBI database. Presence of potential plasmids was checked using PlasmidFinder v.2.1 (http://cge.cbs.dtu.dk/services/PlasmidFinder/, [5]). Identified contigs of potential plasmids were compared to *S. aureus* plasmid p18809-P04 (NC_018968.1) and visualised using BRIG v.0.95 [6]. PHASTER (https://phaster.ca, [7]) was used to identify presence of phages.

**Identification of acquired antibiotic resistance genes and chromosomal mutations.**

SRST2 v0.2.0 [4] with ARGannot database (https://github.com/katholt/srst2/tree/master/data) to identify acquired antimicrobial resistance genes. Chromosomal mutations associated with antibiotic resistance in *gyrA, grlA, grlB* (specific mutations confer resistance to ciprofloxacin), *fusA* (fusidic acid), *rpoB* (rifampin) and *dfrB* (trimethoprim) [8]; as well as additional genes *pbp2*, *pbp4*, *ileS* and *23S* were checked using PointFinder database (last update: 2019-07-02; https://bitbucket.org/genomicepidemiology/pointfinder_db/src/master/)

using ResFinder 4.0 [9] available on Center for Genomic Epidemiology website (http://www.genomicepidemiology.org).

**Genetic relatedness of six *S. argenteus* strains.** Pair-wise Single Nucleotide Polymorphisms (SNPs) were identified using Snippy v3.0 (https://github.com/tseemann/snippy) and Sarg_A1 assembly as reference and SNPs matrix was build using MEGAX v.10.2.4 [10].

**Phylogenetic analysis and comparison to global *S. argenteus* strains.** All available *S. argenteus* genomes submitted to NCBI were downloaded (first downloaded on 07/08/2020, n=129; double checked on 07/02/2021, n=167) and characterised by determining MLST to select ST2250 genomes for comparative analysis. A total of 118 genomes of ST2250 *S. argenteus* were identified from global collection (Suppl. Table S5). SNPs for all ST2250 *S. argenteus* genomes were called using Snippy v3.0 (https://github.com/tseemann/snippy). Maximum-likelihood phylogenetic reconstruction of whole genome alignment was done using IQ-TREE v.2.0.3 [11] with best fit model "K3Pu+F+R4" (determined through extended model selection implemented in IQ-TREE) and 1000 bootstrap replicates. Phylogenetic tree was visualised using iTOL v.5 [12]. SNP pairwise distances were calculated using MEGA X v.10.2.4 [10]. Pangenome of ST2250 was analysed using Roary v.3.13.0 [13].

### Supplementary Tables

**Table S1. Accession numbers and assembly summary of six *S. argenteus* isolates.**

| Isolate ID | Accession no. | Genome length | No. of contigs* | N50 |
|---|---|---|---|---|
| Sarg_A1 | ERS5455722 (SAMEA7699208) | 2745379 | 28 | 244106 |
| Sarg_B1 | ERS5455723 (SAMEA7699209) | 2744813 | 27 | 249010 |
| Sarg_C1 | ERS5455724 (SAMEA7699210) | 2745030 | 22 | 393718 |
| Sarg_C2 | ERS5455725 (SAMEA7699211) | 2743389 | 40 | 168063 |
| Sarg_D1 | ERS5455726 (SAMEA7699212) | 2744155 | 32 | 217193 |
| Sarg_D2 | ERS5455727 (SAMEA7699213) | 2744075 | 44 | 169517 |

*number of contigs longer than 500bp

**Table S2. Summary of Royal Marine recruits carrying *S. argenteus*.** *S. argenteus* isolates are indicated in bold, the other *spa-type* and CC indicated for co-carried *S. aureus*.

| Recruit | Week 1 | Week 6 | Week 15 | Week 32 | Age range (yrs) | SSTI |
|---|---|---|---|---|---|---|
| **A** | t008(CC8)N | **t5078 (CC75)T**<br>t316(CC59)N<br>t1504(CC30)N | t015(CC45)T | x | 16-20 | No |
| **B** | x | x | **t5078 (CC75)T**<br>t015(CC45)T | x | 21-25 | No |
| **C** | x | **t5078(CC75)N**<br>**t5078(CC75)T** | x | x | 16-20 | Yes |
| **D** | **t5078 (CC75)N**<br>**t5078 (CC75)T**<br>t6605(CC398)T | x | t189(CC1)N | x | 21-25 | No |

N – nose swab; T – throat swab; CC- clonal complex was inferred from spa-type; x – no carriage was found, or swab sample was not available.

**Table S3. Chromosomal mutations identified in genes associated with antimicrobial resistance.** None of the identified mutations were previously reported to lead to non-susceptibility to a particular antibiotic. All six *S. argenteus* strains had identical mutations identified in the genes presented in the table.

| | dfrB | pbp2 | gyrA | ileS | grlB | grlA | rpoB | pbp4 | fusA | 23S |
|---|---|---|---|---|---|---|---|---|---|---|
| Sarg_A1 Sarg_B1 Sarg_C1 Sarg_C2 Sarg_D1 Sarg_D2 | no gene | E487D, A462T, D489E, N727_None2181del, A576S, N251D, N472S, A606D, A285K, 711_None712insNS, N386D, R717N, I725L, E319D, D270E, L722I, D430E, S595T, N709S, E269Q, A480S, F646Y, Q629P | S883A, E862D, T827A, N860T, V248E, T818A, V623I, V406I, D856E, D612E, R837H, Q836R | Y905L, R903T, V883G, L898A, N885K, S878I, D276G, M900G, G890W, A223S, I892Y, N901E, E636D, N257D, S877V, Q893S, R894E, L899V, Y288F, V220I, E259Q, *902L, S570A, M873H, E472D, M882D, S546R, I904H, K911E, L912V, N429S, D313E, L870N, *913V, V886C, V889C, K884E, Q881A, K887E, A909C, H907P, V874G, T289S, I263V, M880H,868_None2602del, N879E, N872E, I895D, M871Y, L876I, A906C, D866_None2598del, Q421K, Q869T, D888R, N910Q, D665E, I669V, V897G, I875D, T891N, D908R, N213D, A280D, H915S, N914K | no gene | no gene | I831V, I315L, G225S, | no gene | S238A | 131G>A, 1550G>A, |

**Table S4. Pair-wise SNPs differences of the six *S. argenteus* isolates characterised in this study.**

| | Sarg_B1 | Sarg_C1 | Sarg_C2 | Sarg_D1 | Sarg_D2 | Sarg_A1 |
|---|---|---|---|---|---|---|
| Sarg_B1 | | | | | | |
| Sarg_C1 | 18 | | | | | |
| Sarg_C2 | 17 | 3 | | | | |
| Sarg_D1 | 12 | 18 | 17 | | | |
| Sarg_D2 | 13 | 11 | 10 | 15 | | |
| Sarg_A1 | 22 | 34 | 35 | 30 | 33 | |

**Table S5. Assemblies of *S. argenteus* ST2250 downloaded from NCBI database, relevant publications; and used for comparative analysis in this study.**

| No. | Strain | MLST | Country (reference) | Year | BioSample | BioProject | Assembly |
|---|---|---|---|---|---|---|---|
| 1 | 58113 | ST2250 | Japan [14] | | SAMD00114619 | PRJDB6867 | GCA_003967115.1 |
| 2 | XNO62 | ST2250 | | | SAMN07510741 | PRJNA398564 | GCA_003595345.1 |
| 3 | XNO106 | ST2250 | | | SAMN07510742 | PRJNA400109 | GCA_002812345.2 |
| 4 | 3688STDY6125080 | ST2250 | Thailand [15] | 2013 | SAMEA3449088 | PRJEB9575 | GCA_900126665.1 |

| | | | | | | | |
|---|---|---|---|---|---|---|---|
| 5 | 3688STDY6125091 | ST2250 | Thailand [15] | 2012 | SAMEA3449099 | PRJEB9575 | GCA_900128265.1 |
| 6 | 3688STDY6125064 | ST2250 | Thailand [15] | 2011 | SAMEA3449072 | PRJEB9575 | GCA_900126555.1 |
| 7 | 3688STDY6125066 | ST2250 | Thailand [15] | 2011 | SAMEA3449074 | PRJEB9575 | GCA_900126565.1 |
| 8 | 3688STDY6125116 | ST2250 | Thailand [15] | 2012 | SAMEA3449118 | PRJEB9575 | GCA_900126875.1 |
| 9 | 3688STDY6125100 | ST2250 | Thailand [15] | 2013 | SAMEA3449102 | PRJEB9575 | GCA_900126745.1 |
| 10 | 3688STDY6125126 | ST2250 | Thailand [15] | 2013 | SAMEA3449128 | PRJEB9575 | GCA_900126945.1 |
| 11 | 3688STDY6125111 | ST2250 | Thailand [15] | 2011 | SAMEA3449113 | PRJEB9575 | GCA_900126825.1 |
| 12 | 3688STDY6125070 | ST2250 | Thailand [15] | 2012 | SAMEA3449078 | PRJEB9575 | GCA_900126585.1 |
| 13 | 3688STDY6125063 | ST2250 | Thailand [15] | 2011 | SAMEA3449071 | PRJEB9575 | GCA_900126525.1 |
| 14 | 3688STDY6125068 | ST2250 | Thailand [15] | 2011 | SAMEA3449076 | PRJEB9575 | GCA_900128165.1 |
| 15 | 3688STDY6125072 | ST2250 | Thailand [15] | 2012 | SAMEA3449080 | PRJEB9575 | GCA_900126605.1 |
| 16 | 3688STDY6125075 | ST2250 | Thailand [15] | 2012 | SAMEA3449083 | PRJEB9575 | GCA_900128195.1 |
| 17 | 3688STDY6125077 | ST2250 | Thailand [15] | 2013 | SAMEA3449085 | PRJEB9575 | GCA_900128205.1 |
| 18 | 3688STDY6125105 | ST2250 | Thailand [15] | 2012 | SAMEA3449107 | PRJEB9575 | GCA_900126775.1 |
| 19 | 3688STDY6125121 | ST2250 | Thailand [15] | 2012 | SAMEA3449123 | PRJEB9575 | GCA_900126915.1 |
| 20 | 3688STDY6125113 | ST2250 | Thailand [15] | 2011 | SAMEA3449115 | PRJEB9575 | GCA_900128325.1 |
| 21 | 3688STDY6125093 | ST2250 | | | SAMEA3449101 | PRJEB9575 | GCA_900126735.1 |
| 22 | 3688STDY6125084 | ST2250 | Thailand [15] | 2013 | SAMEA3449092 | PRJEB9575 | GCA_900126695.1 |
| 23 | 3688STDY6125089 | ST2250 | Thailand [15] | 2013 | SAMEA3449097 | PRJEB9575 | GCA_900128255.1 |
| 24 | 3688STDY6125119 | ST2250 | Thailand [15] | 2012 | SAMEA3449121 | PRJEB9575 | GCA_900126895.1 |
| 25 | 3688STDY6125071 | ST2250 | Thailand [15] | 2012 | SAMEA3449079 | PRJEB9575 | GCA_900126595.1 |
| 26 | 3688STDY6125076 | ST2250 | Thailand [15] | 2012 | SAMEA3449084 | PRJEB9575 | GCA_900126625.1 |
| 27 | 3688STDY6125085 | ST2250 | Thailand [15] | 2013 | SAMEA3449093 | PRJEB9575 | GCA_900128225.1 |
| 28 | 3688STDY6125138 | ST2250 | Thailand [15] | 2007 | SAMEA3449140 | PRJEB9575 | GCA_900127075.1 |
| 29 | 3688STDY6125069 | ST2250 | Thailand [15] | 2012 | SAMEA3449077 | PRJEB9575 | GCA_900128175.1 |
| 30 | 3688STDY6125087 | ST2250 | Thailand [15] | 2013 | SAMEA3449095 | PRJEB9575 | GCA_900126705.1 |
| 31 | 3688STDY6125083 | ST2250 | Thailand [15] | 2013 | SAMEA3449091 | PRJEB9575 | GCA_900126685.1 |
| 32 | 3688STDY6125106 | ST2250 | Thailand [15] | 2012 | SAMEA3449108 | PRJEB9575 | GCA_900126785.1 |
| 33 | 3688STDY6125090 | ST2250 | Thailand [15] | 2013 | SAMEA3449098 | PRJEB9575 | GCA_900126725.1 |
| 34 | 3688STDY6125122 | ST2250 | Thailand [15] | 2013 | SAMEA3449124 | PRJEB9575 | GCA_900128345.1 |
| 35 | 3688STDY6125078 | ST2250 | Thailand [15] | 2013 | SAMEA3449086 | PRJEB9575 | GCA_900126645.1 |
| 36 | 3688STDY6125088 | ST2250 | Thailand [15] | 2013 | SAMEA3449096 | PRJEB9575 | GCA_900128245.1 |
| 37 | 3688STDY6125136 | ST2250 | Thailand [15] | 2007 | SAMEA3449138 | PRJEB9575 | GCA_900127045.1 |
| 38 | 3688STDY6125125 | ST2250 | Thailand [15] | 2013 | SAMEA3449127 | PRJEB9575 | GCA_900126935.1 |
| 39 | 3688STDY6125086 | ST2250 | Thailand [15] | 2013 | SAMEA3449094 | PRJEB9575 | GCA_900128235.1 |
| 40 | 3688STDY6125092 | ST2250SLV | Thailand [15] | 2012 | SAMEA3449100 | PRJEB9575 | GCA_900128275.1 |
| 41 | 3688STDY6125140 | ST2250 | Thailand [15] | 2011 | SAMEA3449142 | PRJEB9575 | GCA_900127085.1 |
| 42 | 3688STDY6125114 | ST2250 | Thailand [15] | 2012 | SAMEA3449116 | PRJEB9575 | GCA_900128335.1 |
| 43 | 3688STDY6125062 | ST2250 | Thailand [15] | 2011 | SAMEA3449070 | PRJEB9575 | GCA_900128145.1 |
| 44 | 3688STDY6125112 | ST2250 | Thailand [15] | 2011 | SAMEA3449114 | PRJEB9575 | GCA_900126835.1 |
| 45 | 3688STDY6125115 | ST2250 | Thailand [15] | 2012 | SAMEA3449117 | PRJEB9575 | GCA_900126855.1 |
| 46 | 3688STDY6125081 | ST2250 | Thailand [15] | 2013 | SAMEA3449089 | PRJEB9575 | GCA_900126675.1 |

| | | | | | | | |
|---|---|---|---|---|---|---|---|
| 47 | 3688STDY6125124 | ST2250 | Thailand [15] | 2013 | SAMEA3449126 | PRJEB9575 | GCA_900128365.1 |
| 48 | 3688STDY6125110 | ST2250 | Thailand [15] | 2011 | SAMEA3449112 | PRJEB9575 | GCA_900126815.1 |
| 49 | 3688STDY6125065 | ST2250 | Thailand [15] | 2011 | SAMEA3449073 | PRJEB9575 | GCA_900126545.1 |
| 50 | 3688STDY6125067 | ST2250 | Thailand [15] | 2011 | SAMEA3449075 | PRJEB9575 | GCA_900128155.1 |
| 51 | 3688STDY6125082 | ST2250 | Thailand [15] | 2013 | SAMEA3449090 | PRJEB9575 | GCA_900128215.1 |
| 52 | 3688STDY6125108 | ST2250 | Thailand [15] | 2013 | SAMEA3449110 | PRJEB9575 | GCA_900126795.1 |
| 53 | 3688STDY6125073 | ST2250 | Thailand [15] | 2012 | SAMEA3449081 | PRJEB9575 | GCA_900128185.1 |
| 54 | 3688STDY6125131 | ST2250 | Thailand [15] | 2006 | SAMEA3449133 | PRJEB9575 | GCA_900126985.1 |
| 55 | 3688STDY6125118 | ST2250 | Thailand [15] | 2012 | SAMEA3449120 | PRJEB9575 | GCA_900126865.1 |
| 56 | 3688STDY6125109 | ST2250 | Thailand [15] | 2011 | SAMEA3449111 | PRJEB9575 | GCA_900126805.1 |
| 57 | 3688STDY6125120 | ST2250 | Thailand [15] | 2012 | SAMEA3449122 | PRJEB9575 | GCA_900126905.1 |
| 58 | 3688STDY6125123 | ST2250 | Thailand [15] | 2013 | SAMEA3449125 | PRJEB9575 | GCA_900128355.1 |
| 59 | 3688STDY6125074 | ST2250 | Thailand [15] | 2012 | SAMEA3449082 | PRJEB9575 | GCA_900126615.1 |
| 60 | 3688STDY6125117 | ST2250 | Thailand [15] | 2012 | SAMEA3449119 | PRJEB9575 | GCA_900126885.1 |
| 61 | LBSA043 | ST2250 | | | SAMEA2007935 | PRJEB6393 | GCA_000752055.1 |
| 62 | SCPM-O-B-8378 | ST2250 | | | SAMN08689422 | PRJNA269675 | GCA_003011845.1 |
| 63 | ST2250 | ST2250 | | | SAMN09435802 | PRJNA476500 | GCA_003935695.1 |
| 64 | 3688STDY6125079 | ST2250 | Thailand [15] | 2013 | SAMEA3449087 | PRJEB9575 | GCA_900126655.1 |
| 65 | PHL3431 | ST2250 | | | SAMN09704415 | PRJNA482405 | GCA_009874245.1 |
| 66 | SARG0275 | ST2250 | | | SAMD00133958 | PRJDB7256 | GCA_005397745.1 |
| 67 | PHL3433 | ST2250 | | | SAMN09704514 | PRJNA482411 | GCA_008368955.1 |
| 68 | RK308 | ST2250 | | | SAMN04457463 | PRJNA310972 | GCA_001641625.1 |
| 69 | H2179 | ST2250 | Denmark [16] | 2013 | SAMEA104034175 | PRJEB20633 | GCA_900183785.1 |
| 70 | M4611 | ST2250 | Denmark [16] | 2013 | SAMEA104034174 | PRJEB20633 | GCA_900183845.1 |
| 71 | M3040 | ST2250 | Denmark [16] | 2016 | SAMEA104034155 | PRJEB20633 | GCA_900183955.1 |
| 72 | CCUG 69384 | ST2250 | Sweden [17] | 2016 | SAMN07566689 | PRJNA305687 | GCA_003010635.1 |
| 73 | M21126 | ST2250 | | | SAMN02402619 | PRJNA227695 | GCA_000606605.1 |
| 74 | M4143 | ST2250 | Denmark [16] | 2013 | SAMEA104034168 | PRJEB20633 | GCA_900183925.1 |
| 75 | SJTU F21285 | ST2250 | | | SAMN04604746 | PRJNA317282 | GCA_001969725.1 |
| 76 | F87619 | ST2250 | | | SAMN02402601 | PRJNA227677 | GCA_000602805.1 |
| 77 | 1299_SAUR | ST2250 | | | SAMN03197271 | PRJNA267549 | GCA_001061465.1 |
| 78 | O-6 | ST2250 | Denmark [16] | 2015 | SAMEA104034161 | PRJEB20633 | GCA_900183945.1 |
| 79 | SJTU F21224 | ST2250 | | | SAMN04604745 | PRJNA317280 | GCA_001969715.1 |
| 80 | MGYG-HGUT-02548 | ST2250 | | | SAMEA5852053 | PRJEB33885 | GCA_902388265.1 |
| 81 | H1826 | ST2250 | Denmark [16] | 2014 | SAMEA104034166 | PRJEB20633 | GCA_900183825.1 |
| 82 | H1540 | ST2250 | Denmark [16] | 2014 | SAMEA104034164 | PRJEB20633 | GCA_900183815.1 |
| 83 | O-1 | ST2250 | Denmark [16] | 2016 | SAMEA104034156 | PRJEB20633 | GCA_900183865.1 |
| 84 | SJTU F20419 | ST2250 | | | SAMN04604743 | PRJNA317278 | GCA_001969695.1 |
| 85 | O-5 | ST2250 | Denmark [16] | 2015 | SAMEA104034160 | PRJEB20633 | GCA_900183895.1 |
| 86 | O-2 | ST2250SLV | Denmark [16] | 2016 | SAMEA104034157 | PRJEB20633 | GCA_900183965.1 |
| 87 | O-3 | ST2250 | Denmark [16] | 2016 | SAMEA104034158 | PRJEB20633 | GCA_900183765.1 |

| | | | | | | | |
|---|---|---|---|---|---|---|---|
| 88 | 5506 | ST2250 | | | SAMN13979052 | PRJNA595347 | GCA_010570875.1 |
| 89 | SJTU F21164 | ST2250 | | | SAMN04604744 | PRJNA317279 | GCA_001969705.1 |
| 90 | O-4 | ST2250 | Denmark [16] | 2015 | SAMEA104034159 | PRJEB20633 | GCA_900183795.1 |
| 91 | M4148 | ST2250 | Denmark [16] | 2013 | SAMEA104034170 | PRJEB20633 | GCA_900183875.1 |
| 92 | M4146 | ST2250 | Denmark [16] | 2013 | SAMEA104034169 | PRJEB20633 | GCA_900183755.1 |
| 93 | H1604 | ST2250 | Denmark [16] | 2014 | SAMEA104034165 | PRJEB20633 | GCA_900183775.1 |
| 94 | O-10 | ST2250 | Denmark [16] | 2015 | SAMEA104034163 | PRJEB20633 | GCA_900183855.1 |
| 95 | PFESA3024 | ST2250 | Singapore [15] | 2011 | SAMEA2662162 | PRJEB1915 | GCA_900128685.1 |
| 96 | PFESA2861 | ST2250 | Singapore [15] | 2009 | SAMEA2662140 | PRJEB1915 | GCA_900128635.1 |
| 97 | PFESA2947 | ST2250 | Malaysia [15] | 2011 | SAMEA2661974 | PRJEB1915 | GCA_900128625.1 |
| 98 | PFESA2956 | ST2250 | Malaysia [15] | 2011 | SAMEA2662160 | PRJEB1915 | GCA_900128615.1 |
| 99 | PFESA2944 | ST2250 | Malaysia [15] | 2011 | SAMEA2662156 | PRJEB1915 | GCA_900128705.1 |
| 100 | PFESA2941 | ST2250 | Malaysia [15] | 2011 | SAMEA2662088 | PRJEB1915 | GCA_900128645.1 |
| 101 | PFESA2952 | ST2250 | Malaysia [15] | 2011 | SAMEA2662072 | PRJEB1915 | GCA_900128655.1 |
| 102 | PFESA2963 | ST2250 | Malaysia [15] | 2011 | SAMEA2662127 | PRJEB1915 | GCA_900128665.1 |
| 103 | PFESA3295 | ST2250 | Israel [15] | 2009 | SAMEA2710365 | PRJEB1915 | GCA_900128605.1 |
| 104 | PFESA2045 | ST2250 | France [15] | 2010 | SAMEA2445518 | PRJEB1915 | GCA_900128675.1 |
| 105 | T151204 | ST2250 | France [18] | | SAMEA6530084 | PRJEB36681 | |
| 106 | MC3 | ST2250 | Canada [19] | | SAMN16304021 | PRJNA666697 | GCA_014877945.1 |
| 107 | WU2 | ST2250 | Canada [19] | | SAMN16304024 | PRJNA666697 | GCA_014877415.1 |
| 108 | MC4 | ST2250 | Canada [19] | | SAMN16304022 | PRJNA666697 | GCA_014877235.1 |
| 109 | MC2 | ST2250 | Canada [19] | | SAMN16304019 | PRJNA666697 | GCA_014877005.1 |
| 110 | WU1 | ST2250 | Canada [19] | | SAMN16304023 | PRJNA666697 | GCA_014877475.1 |
| 111 | WU3 | ST2250 | Canada [19] | | SAMN16304020 | PRJNA666697 | GCA_014877915.1 |
| 112 | PHL2420 | ST2250 | Canada [19] | | SAMN16304007 | PRJNA666697 | GCA_014878065.1 |
| 113 | PHL4815 | ST2250 | Canada [19] | | SAMN16304012 | PRJNA666697 | GCA_014877035.1 |
| 114 | PHL3446 | ST2250 | Canada [19] | | SAMN16304006 | PRJNA666697 | GCA_014878035.1 |
| 115 | PHL6318 | ST2250 | Canada [19] | | SAMN16304008 | PRJNA666697 | GCA_014878055.1 |
| 116 | PHL6344 | ST2250 | Canada [19] | | SAMN16304004 | PRJNA666697 | GCA_014878095.1 |
| 117 | PHL8605 | ST2250 | Canada [19] | | SAMN16304009 | PRJNA666697 | GCA_014877525.1 |
| 118 | PHL4226 | ST2250 | Canada [19] | | SAMN16304014 | PRJNA666697 | GCA_014877505.1 |

### Supplementary Figures

**Figure S1. Putative plasmid identified in this study from all six *S. argenteus* strains.**

BRIG [6] was used to present BLASTN comparisons of the whole contig of putative plasmid of *S. argenteus* identified in this study and compared. The innermost ring shows genome scale in kilobase pairs and subsequent rings show BLASTN comparisons. **(a)** The innermost ring is putative plasmid contig of isolate Sarg_A1 which is compared to the putative plasmid contigs of other isolates in the outermost rings as follows: (1) Sarg_B1, (2) Sarg_C1, (3) Sarg_C2, (4) Sarg_D1, (5) Sarg_D2 and (6) annotated genes on a Sarg_A1 putative plasmid; non-annotated genes present hypothetical genes. **(b)** NCBI database search reported *S. aureus* plasmid p18809-P04 (NC_018968.1) being the closest match, but large portion of the plasmid was missing.

**(a)**

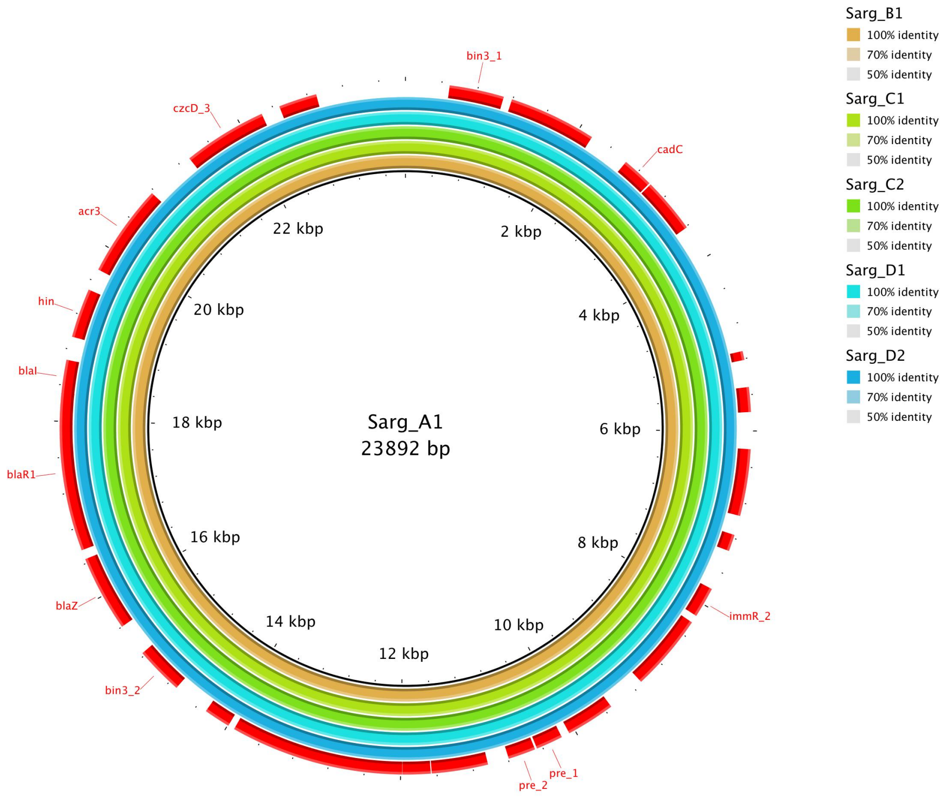

**(b)**

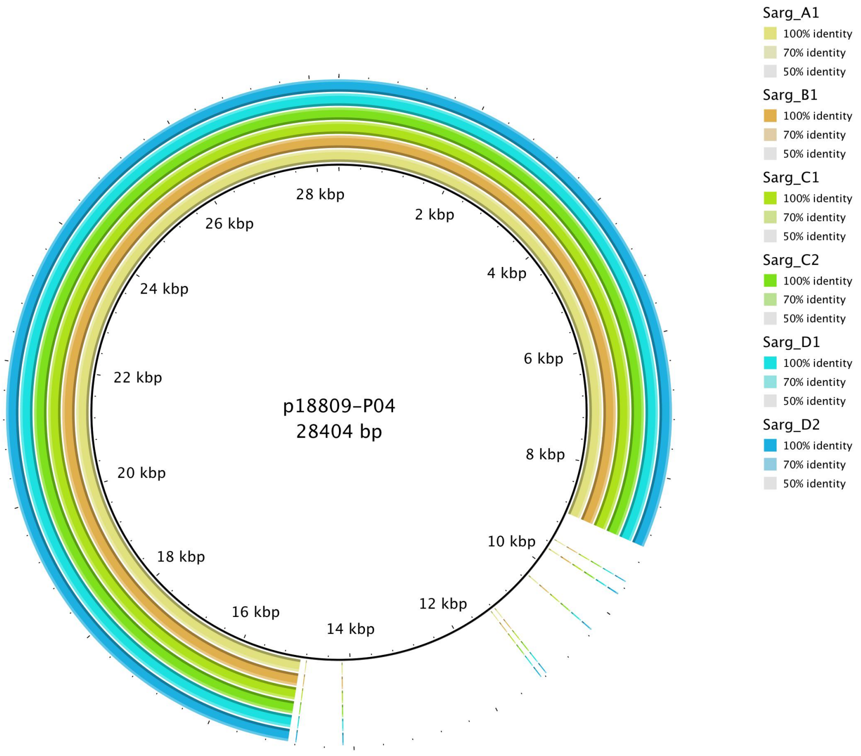

p18809-P04
28404 bp
28 kbp
2 kbp
4 kbp
6 kbp
8 kbp
10 kbp
12 kbp
14 kbp
16 kbp
18 kbp
20 kbp
22 kbp
24 kbp
26 kbp
Sarg_A1
100% identity
70% identity
50% identity
Sarg_B1
100% identity
70% identity
50% identity
Sarg_C1
100% identity
70% identity
50% identity
Sarg_C2
100% identity
70% identity
50% identity
Sarg_D1
100% identity
70% identity
50% identity
Sarg_D2
100% identity
70% identity
50% identity

**Suppl. Material references:**

[9] Bortolaia V, Kaas RS, Ruppe E, Roberts MC, Schwarz S, Cattoir V, et al. ResFinder

4.0 for predictions of phenotypes from genotypes. J Antimicrob Chemother 2020;75. https://doi.org/10.1093/jac/dkaa345.
